# B cell depletion attenuates CD27 signaling of T helper cells in multiple sclerosis

**DOI:** 10.1101/2022.10.17.22281079

**Authors:** Can Ulutekin, Edoardo Galli, Mohsen Khademi, Ilaria Callegari, Fredrik Piehl, Nicholas Sanderson, Massimo Filippi, Roberto Furlan, Tomas Olsson, Tobias Derfuss, Florian Ingelfinger, Burkhard Becher

## Abstract

Multiple sclerosis (MS) is a chronic inflammatory disease of the central nervous system (CNS). Traditionally, MS was held to be a T-cell mediated disease, but accumulating evidence during the last decade also highlighted the crucial importance of B cells for the disease progression. Particularly, B cell depleting therapies (BCDTs), have demonstrated striking efficacy in suppressing inflammatory disease activity in relapsing-remitting MS. However, a detailed understanding of the role of B cells in the pathogenesis of MS is still lacking, and by extension also the mechanism of action of BCDTs. In this longitudinal multi-center study, we investigated the impact of BCDTs on the immune landscape in MS patients using high-dimensional single-cell immunophenotyping (cytometry by time-of-flight; CyTOF). Algorithm-guided analyses revealed phenotypic changes in the newly reconstituted B cell compartment after BCDT, as well as a marked specific reduction of circulating T follicular helper (Tfh) cells with a concomitant upregulation of CD27 surface expression in memory T helper cells and Tfh cells. These findings indicate a costimulatory mechanism in the CD27/CD70 signaling pathway, through which B cells sustain the activation of pathogenic T cells. Disrupting the CD27/CD70 signaling axis via BCDTs provides a potential explanation for its clinical efficacy.

**One Sentence Summary:** B cell depletion contracts follicular T helper cells, displaces memory-to-naïve ratio and impairs CD27 signaling in T helper cells.

## INTRODUCTION

MS is an immune-mediated disease, characterized by demyelination of the CNS, where disease progression results from a complex interplay between partially understood neurodegenerative processes and inflammatory episodes. Traditionally, MS was considered a T cell-mediated disease based on evidence inferred from animal models of neuroinflammation *(1, 2)*. However, therapies targeting exclusively T lymphocytes in patients demonstrated only weak clinical efficacy *(3)*. More recent evidence supported the involvement of B cells in the disease pathogenesis and progression of MS. One of the most convincing findings highlighting the critical role of B cells in the disease has been the high response rate of BCDTs in ameliorating MS *(4–6)*. These findings have shifted the perception of the disease towards mechanisms in which the interplay between B and T cells takes a more prominent role *(7)*.

Despite the impressive clinical efficacy of BCDTs in suppressing inflammatory disease activity in MS, their precise mechanism of action remains unclear and is an area of active research *(8)*. Removal of the total circulating immunoglobulin fraction via plasmapheresis *(9, 10)*, or interfering with potentially pathogenic antibodies via intravenous immune globulin (IVIG) injections *(11–13)* have shown limited efficacy. Furthermore, anti-CD20-mediated BCDTs do not target plasma cells and have minimal effects on antibody titters *(6)*. Thus, it appears likely that the success of BCDTs depends on B-cell activities other than antibody production, such as cytokine secretion, antigen presentation or co-stimulation. Several studies report upregulation of proinflammatory cytokines and a concomitant downregulation of immunoregulatory cytokines in B cells of MS *(14–20)*. Apart from cytokine production, B cells are antigen-presenting cells (APCs) and can efficiently prime T cells *(21)*. Accordingly, polymorphisms in major histocompatibility complex (MHC) genes demonstrate the strongest association with MS susceptibility, particularly those encoding for class II MHC molecules *(22–25)*.

The characterization of the replenished B cell population after depletion with BCDTs has been examined before *(14, 16, 19)*, but studies investigating the immune compartment as a whole are missing. Here, we systematically investigated the effects of BCDTs on the peripheral immune landscape in MS patients. We used high-dimensional mass cytometry in conjunction with unsupervised clustering to analyze the systemic immune compartment of MS patients after BCDTs in a longitudinal manner. B cell depletion evoked alterations in CD4^+^ T cell composition and expression of costimulatory molecules suggesting that the cell-to-cell interactions between B and T cells play a key role during MS immunopathology that could be compromised using BCDTs. Our study thereby addresses the enigmatic role of B cells in MS pathology and suggests a costimulatory mechanism of action for B cell depleting agents in MS patients.

## RESULTS

### The replenished B cell compartment is phenotypically altered in MS

To characterize the immune landscape of MS patients after BCDT, we collected longitudinal peripheral blood mononuclear cell (PBMC) samples of MS patients undergoing BCDT (table 1). The samples were analyzed by two partially overlapping mass cytometry panels in order to deeply interrogate cellular states and subsets associated with the clinical success of BCDT. Dimensionality reduction using Uniform Manifold Approximation and Projection (UMAP) and unsupervised FlowSOM clustering identified B cells, CD4^+^ T cells, CD8^+^ T cells, regulatory T (Treg) cells, γδ T cells, natural killer (NK) cells and myeloid cells (fig. 1, A and B).

**Table 1:** Clinical and demographic characteristics of patients in the discovery cohort.

| ID | Diagnosis | Age Range | Sex | Treatment | Previous Treatment | Cycle of Treatment | Days Since Last Treatment | Days Since First Treatment |
| --- | --- | --- | --- | --- | --- | --- | --- | --- |
| 1 | RRMS | 31 - 40 | M | Ocrelizumab | None | 2 | 146 | 160 |
| 2 | RRMS | 31 - 40 | F | Ocrelizumab | None | 2 | 148 | 162 |
| 3 | RRMS | 21 - 30 | M | Ocrelizumab | Interferon $\beta$ | 2 | 57 | 71 |
| 4 | RRMS | 21 - 30 | F | Ocrelizumab | None | 2 | 43 | 57 |
| 5 | PPMS | 61 - 70 | F | Rituximab | None | 6 | 126 | 877 |
| 6 | PPMS | 51 - 60 | M | Ocrelizumab | None | 3 | 117 | 320 |
| 7 | RRMS | 41 - 50 | F | Ocrelizumab | None | 2 | 43 | 57 |
| 8 | RRMS | 21 - 30 | F | Ocrelizumab | None | 2 | 154 | 168 |
| 9 | RRMS | 31 - 40 | F | Ocrelizumab | None | 2 | 93 | 106 |
| 10 | RRMS | 51 – 60 | F | Ocrelizumab | None | 2 | 182 | 196 |
| 11 | PPMS | 41 – 50 | F | Rituximab | None | 3 | 98 | 306 |
| 12 | RRMS | 51 - 60 | M | Rituximab | None | 3 | 92 | 468 |

**Fig. 1.**
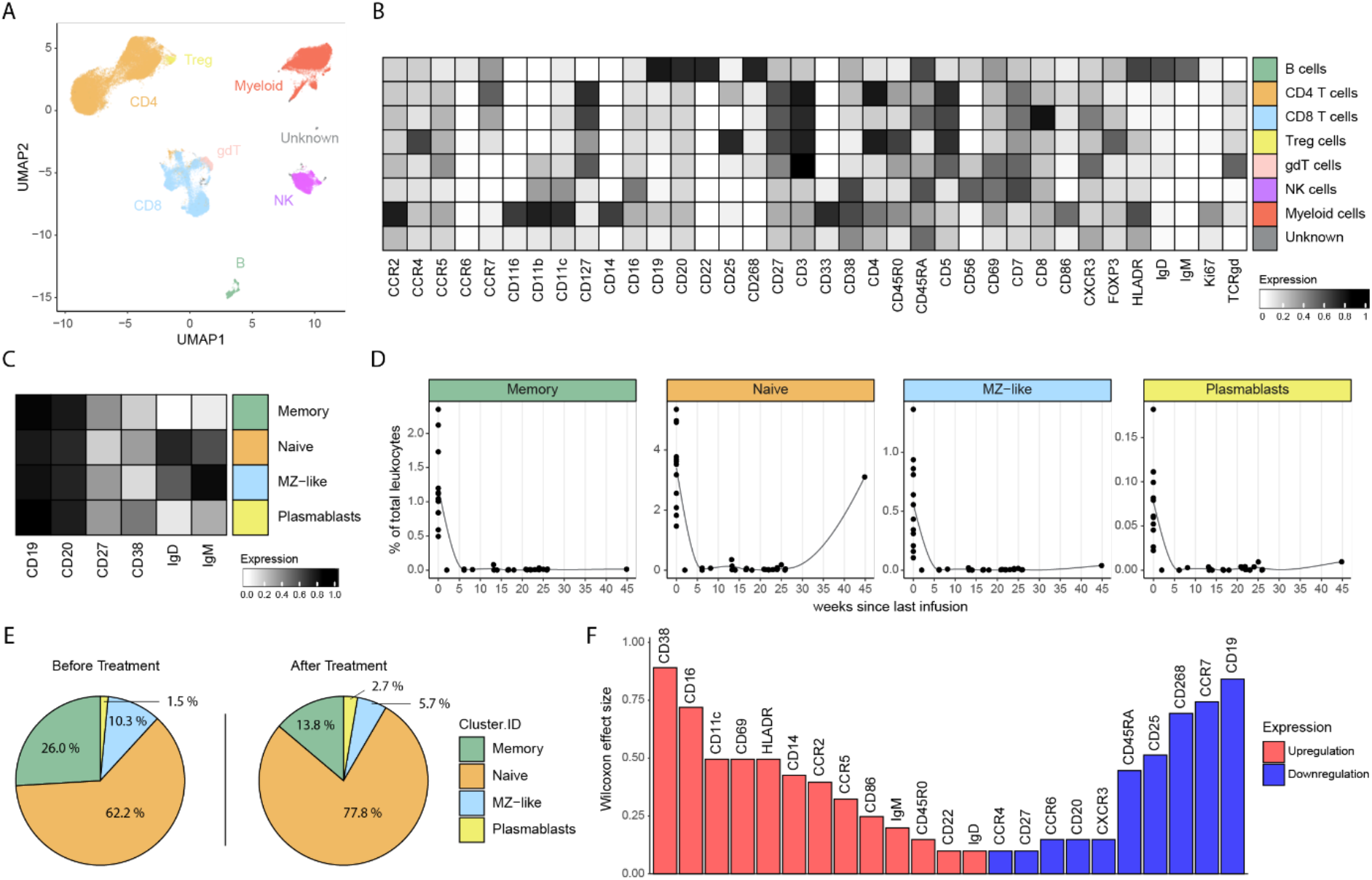
BCDTs deplete B cells efficiently and alter the B cell phenotype after reconstitution. (**A**) UMAP of 52,500 cells randomly sampled from the combined dataset. The number of sampled cells was constant across samples. Color code indicates the main clusters generated through clustering and manual annotation. (**B**) Heatmap showing the median marker expression across the identified main clusters. (**C**) Heatmap shows the median marker expression across the identified B cell sub-clusters. (**D**) Frequency of B cell subsets among total cells across time after infusion. Points represent individual samples. The line represents the conditional mean of B cell subset frequency. (**E**) Pie chart showing the composition of distinct subsets within the B cell compartment before (left) and after (right) treatment. The after-treatment B cell compartment consists of late timepoint representing the replenishing B cell compartment. (**F**) Bar plot displaying changes in marker expression of newly populating B cells in MS. Marker expression changes visualized through paired Wilcoxon effect size. Color code indicates up- or downregulation.

To assess the kinetics of the impact of BCDTs on discrete B cell subsets, B cells were dissected into canonical B cell subsets: memory, naïve, marginal zone-like (MZ-like), and plasmablasts (fig.1C, and fig. S1A). In line with previous reports *(26)*, each of the investigated B cell subsets were efficiently depleted after treatment infusions (fig. 1D). To assess how the newly repopulating B cell compartment differed following BCDT, we compared the B cell compartment before therapy initiation to timepoints, where a replenishment could be observed (8-45 weeks since last infusion, median 22; 106-7041 cell counts, median 399). Consistent with previous reports *(27)* we observed a decrease in the frequency of memory B cells and an increase in the frequency of naïve B cells (fig. 1E).

Newly replenished B cells revealed a downregulation of CCR7, the chemokine receptor essential for migration to lymphoid tissues *(28)* and CD268 and CD19 (fig. 1F and fig. S1B). CD268, encoding for the B-cell-activating receptor (BAFF-R), is a crucial regulator of B- and T-cell activity *(29)*, however, failing evaluation as a pharmacological target in MS *(30)*, in spite of compelling evidence from preclinical studies *(31, 32)*. CD19 is a critical protein of the CD19 complex, involved in the amplification of B cell receptor signaling and subsequent B cell activation *(33, 34)*.

### B cell depletion reduces the frequency of T follicular helper cells in MS patients

To have a broad overview of the impact of BCDTs on the immune compartment, we investigated the frequency of distinct immune cell populations in MS patients before and after therapy. To account for the bias elicited by the depletion of B cells, we computationally removed them from our analysis and normalized the frequencies accordingly. We observed that the canonical immune cell populations were not altered by BCDT treatment suggesting a more nuanced effect of the treatment on the immune landscape (fig. S1C).

B- and T-cell interactions have a fundamental role during the immunopathology of MS *(35)*, and interference with these interactions could partially account for the clinical efficacy of BCDTs. To better characterize the impact of BCDTs on the T cell compartment we defined five distinct CD4^+^ T cell subsets in the peripheral blood of MS patients; central memory, effector memory, naïve, T follicular helper (Tfh) and CD103^+^ cells (fig. 2A). For each subset we longitudinally analyzed the shift in the frequency after BCDT (fig. 2B). We observed a significant decrease in the frequency of Tfh cells in response to BCDT treatment. Circulating Tfh cells have been reported to be increased in the blood of MS patients and a reduction of these has been associated with a remission of the disease *(36, 37)*. These observations suggest that BCDTs may indirectly deplete potentially pathogenic Tfh cells and highlight the interdependence of these with B cells. Consistent with previous observations *(38)*, when comparing changes in the naïve-to-memory ratio, we observed a shift from memory towards naïve Th cells (fig. 2C).

**Fig. 2.**
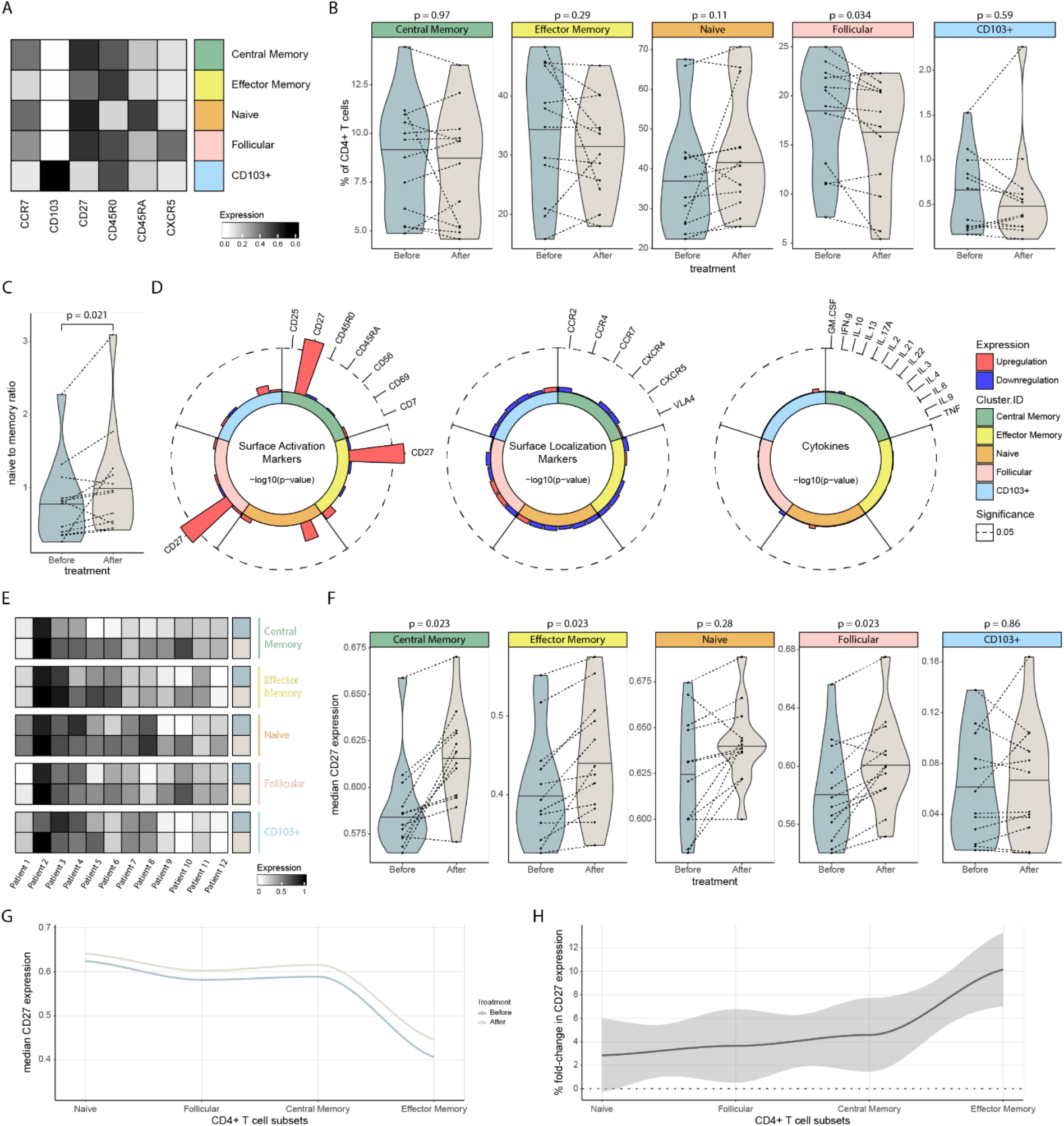
BCDTs alter the peripheral CD4^+^ T cell landscape of MS patients. (**A**) Heatmap showing median expression of lineage markers across identified CD4+ T cell subsets. (**B**) Violin plots showing changes in the frequency across CD4+ T cell subsets in BCDT treated MS. Paired Wilcoxon rank-sum test and Benjamini-Hochberg correction was applied. (**C**) Violin plots showing changes in the naive to memory ratio within the CD4+ T cell compartment. Paired Wilcoxon rank-sum test was applied. (**D**) Radar plots showing changes in the marker expression of CD4+ T cell subsets in BCDT treated MS. Values correspond to -log10(p-value) and were adjusted with the Benjamini-Hochberg correction. Inner circle color annotations denote each respective subset. Bar color denotes the upregulation or downregulation of the respective marker. Marker order is consistent across subsets. (**E**) Various heatmaps showing CD27 expression of each MS patient before and after B cell depleting treatment. Each heatmap denotes a different CD4+ T cell subset. CD27 expression on each heatmap normalized by transforming the range of expression to maximal separation within 0 and 1 values. (**F**) Violin plots showing changes in CD27 expression across CD4+ T cell subsets in BCDT treated MS. P values correspond to the paired Wilcoxon rank-sum test applying a Benjamini-Hochberg correction. (**G**) Median CD27 expression along CD4^+^ T cell subsets of the MS patients. Points are individual sample values. Color code denotes patient values before or after treatment. (**H**) Percent fold-change in CD27 expression along CD4^+^ T cell subsets of the MS patients, depicted in percent changes. Shaded area represents the 95 % confidence interval.

Analogous, to the CD4^+^ T cell compartment, we defined distinct subsets within the CD8^+^ T cell compartment (fig. S2A). Analyzing the shift in the frequency of these subsets in BCDT treated MS patients, we observed that these were largely unaffected in response to BCDT treatment (fig. S2B). Accordingly, the ratio of naïve-to-memory cytotoxic T cells was unchanged between untreated and treated MS (fig. S2C). Despite the dominance of CD8^+^ T cells over CD4^+^ T cells in lesions of MS patients and their potential involvement in the immunopathology of MS *(39–41)*, these results point towards a highly selective influence of BCDTs on the Th cell compartment.

### B cell depletion leads to selective increase in CD27 expression across memory Th cell subsets

Next, we assessed whether phenotypic alterations in CD4^+^ T cell subsets could be associated with BCDTs. To this end, we assigned each marker to one of three distinct, mutually exclusive groups: surface activation markers, surface localization markers and cytokines. For each subset we compared longitudinally how the expression levels changed in response to B cell depletion (fig. 2D). This approach revealed a selective disparity in CD27 expression across T helper (Th) cell subsets, where we observed increased surface CD27 across central and effector memory Th cells as well as Tfh cells (fig. 2, E and F). In relation to our observations, soluble CD27 (sCD27) in the cerebrospinal fluid (CSF) is a prognostic biomarker in MS and its protein levels correlate directly with disease severity *(42–44)*.

The costimulatory molecule CD27 is part of the tumor necrosis factor receptor superfamily (TNFRSF) and its ligand CD70 is expressed by APCs *(45–48)*. CD27 signaling leads to the induction of pro-survival pathways*(49, 50)* and supports the efficient expansion and differentiation of activated T cells *(51–54)*. CD27 expression is transiently upregulated upon TCR-engagement *(55–57)* and proteolytically cleaved and released as soluble CD27 (sCD27) upon sustained signaling *(42, 58, 59)*. Surface expression of CD27 therefore naturally declines along the T cell differentiation path *(47)*, with naïve T cells having the highest expression and more differentiated subsets showing a lower expression. Thus, higher surface CD27 expression levels may be a result of impaired interaction due to the depletion of CD70 expressing B cells and could represent an attenuation of CD27 signaling. Indeed, when we examined surface CD27 levels along the differentiation path of Th cells we observed an overall increased expression after BCDT (fig. 2G). This effect was most pronounced in more differentiated Th cells (fig. 2H). In summary, elevated surface levels of CD27 is likely the consequence of decreased engagement with CD70 on B cells, which could partially hamper the pathogenic potential of Th cells.

Next, we assessed changes in marker expression in the CD8^+^ T cell compartment analogous to the previous analysis for the CD4^+^ T cells. None of the analyzed markers appeared dysregulated after BCDT in the cytotoxic T cell compartment (fig. S2D). Specifically, CD27 expression levels during cytotoxic T cell differentiation followed a similar expression pattern as observed for the CD4^+^ T cell counterpart but did not reveal any changes as a result of BCDT, again highlighting the selectivity of BCDT on Th cell compartment (fig. S2, E and F).

### Analysis of independent cohort confirmed the impact of BCDTs on circulatory Tfh and CD27 expression

Next, we investigated whether the selective changes of BCDT on the Th cell compartment could be validated in an independent cohort derived from a distinct clinical center (table 2). This confirmed the reduction of circulating Tfh cells in response to BCDTs in an additional dataset (fig. 3A). When analyzing changes in CD27 expression across CD4^+^ memory T cell and Tfh cell subsets, we confirmed our previous result and observed significant upregulation in surface CD27 levels in central memory T cells and Tfh cells but not effector memory T cells (fig. 3B). In conclusion, using an independent cohort of BCDT treated MS patients, we confirmed our findings that Tfh cells specifically decrease in frequency and surface CD27 levels of central memory T cells and Tfh cells increase after treatment.

**Table 2:** Clinical and demographic characteristics of MS patients in the validation cohort.

| ID | Diagnosis | Age Range | Sex | Treatment | Previous Treatment | Cycle of Treatment | Days Since Last Treatment | Days Since First Treatment |
| --- | --- | --- | --- | --- | --- | --- | --- | --- |
| 1 | RRMS | 21 – 30 | M | Rituximab | None | 1 | 185 | 185 |
| 2 | RRMS | 31 – 40 | F | Rituximab | None | 1 | 182 | 182 |
| 3 | RRMS | 21 - 30 | F | Rituximab | None | 3 | 112 | 475 |
| 4 | RRMS | 41 – 50 | F | Rituximab | Cortisone | 3 | 91 | 455 |
| 5 | RRMS | 21 – 30 | F | Rituximab | Cortisone | 2 | 188 | 360 |
| 6 | RRMS | 21 – 30 | F | Rituximab | None | 3 | 171 | 534 |
| 7 | RRMS | 21 – 30 | F | Rituximab | None | 3 | 108 | 468 |
| 8 | RRMS | 31 – 40 | F | Rituximab | None | 2 | 234 | 463 |
| 9 | RRMS | 41 – 50 | M | Rituximab | None | 6 | 126 | 1050 |
| 10 | RRMS | 31 – 40 | M | Rituximab | None | 6 | 12 | 925 |
| 11 | RRMS | 51 – 60 | M | Rituximab | None | 5 | 127 | 851 |
| 12 | RRMS | 51 – 60 | F | Rituximab | None | 2 | 27 | 207 |
| 13 | RRMS | 41 – 50 | M | Rituximab | None | 1 | 130 | 130 |
| 14 | RRMS | 31 - 40 | M | Rituximab | Cortisone | 1 | 90 | 90 |

**Fig. 3.**
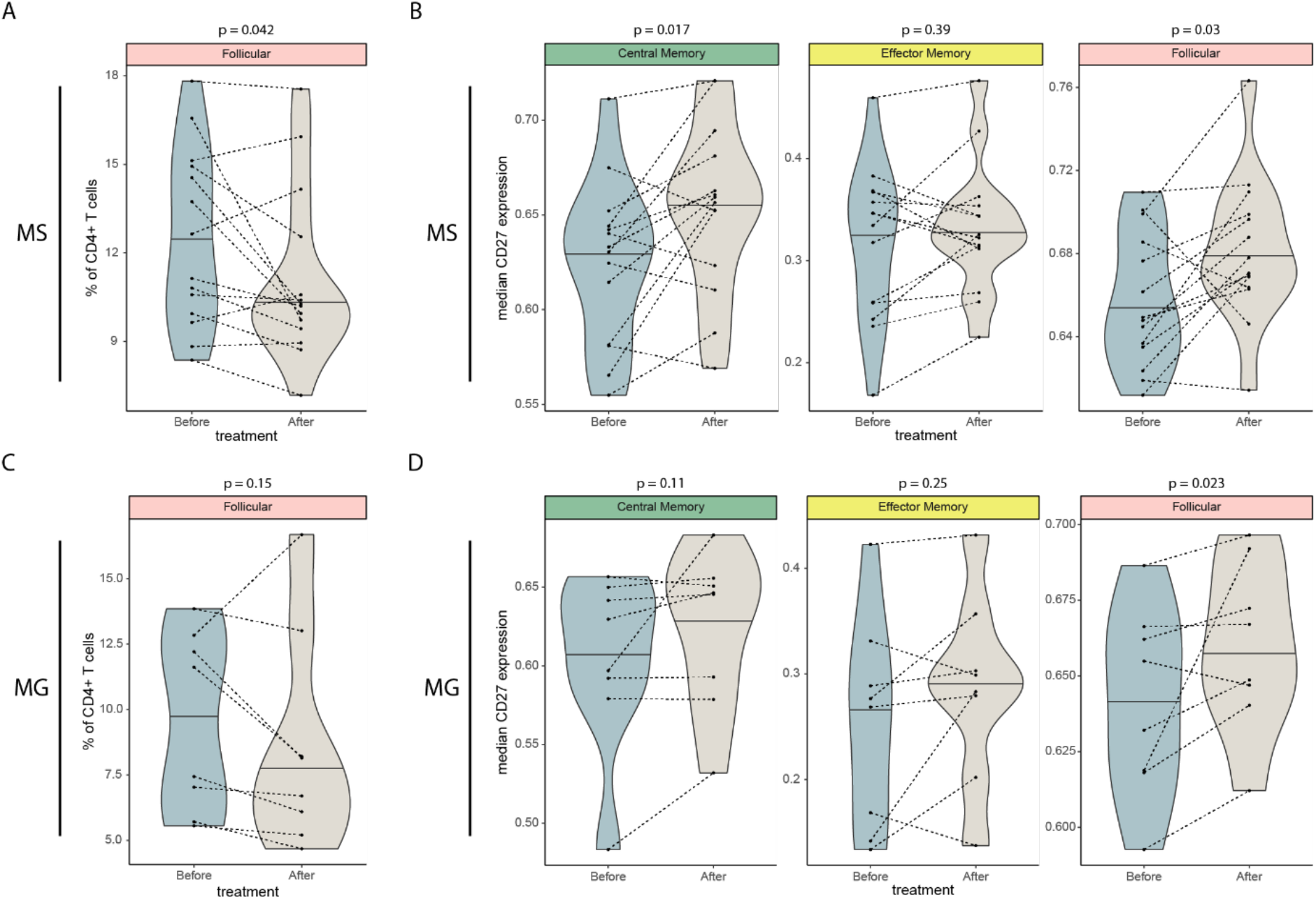
The altered CD4^+^ T cell landscape is consistent across cohorts. (**A**) Violin plot showing the frequency in CD4^+^ Tfh cells in B cell depleted MS patients of the validation cohort. Paired Wilcoxon rank-sum test was applied. (**B**) Violin plots showing changes in CD27 expression across CD4^+^ memory and Tfh cells in B cell depleted MS patients of the validation cohort. Paired Wilcoxon rank-sum test was applied. (**C**) Violin plot showing the frequency in CD4^+^ Tfh cells in B cell depleted MG patients of the validation cohort. Paired Wilcoxon rank-sum test was applied. (**D**) Violin plots showing changes in CD27 expression across CD4^+^ memory and Tfh cells in B cell depleted MG patients of the validation cohort. Paired Wilcoxon rank-sum test was applied.

### BCDT in myasthenia gravis patients induced CD27 expression in Tfh cells

To assess whether the selective influence of BCDTs on the Th cell compartment was specific to MS pathophysiology or a universal feature of BCDTs. We next analyzed the influence of BCDTs on the CD4^+^ T cell compartment in myasthenia gravis (MG) patients in an independent cohort (table 3). For comparative purposes, these patients were selected to be treatment naïve to immune suppressants, except for exposure to low to moderate doses of corticosteroids in some cases. Longitudinal comparison of Tfh cell frequencies revealed a decreasing trend that did not reach statistical significance (fig. 3C). Similarly, we evaluated surface CD27 levels in memory T cells and Tfh cells in response to BCDTs. Consistent with the MS findings, we observed a trend towards upregulation (fig. 3D). However only in Tfh cells this measurement reached statistical significance. The number of MG patients analyzed here is relatively small, thus limiting the statistical power. Despite the disparity in clinical efficacy of BCDTs in MS and MG *(60, 61)*, we can conclude that upregulation of CD27 in Tfh cells is a universal hallmark of the therapy.

**Table 3:** Clinical and demographic characteristics of MG patients in the validation cohort.

| ID | Serology | Age Range | Sex | Treatment | Previous Treatment | Cycle of Treatment | Days Since Last Treatment | Days Since First Treatment |
| --- | --- | --- | --- | --- | --- | --- | --- | --- |
| 15 | LRP4 | 61 - 70 | M | Rituximab | None | 1 | 197 | 197 |
| 16 | AChR | 41 – 50 | M | Rituximab | Mestinon | 1 | 215 | 215 |
| 17 | LRP4 | 51 – 60 | M | Rituximab | Cortisone, Mestinon | 1 | 159 | 159 |
| 18 | AChR | 21 – 30 | F | Rituximab | None | 1 | 91 | 91 |
| 19 | AChR | 41 – 50 | F | Rituximab | None | 1 | 91 | 91 |
| 20 | Negative | 51 – 60 | M | Rituximab | None | 1 | 115 | 115 |
| 21 | Negative | 41 – 50 | F | Rituximab | None | 1 | 226 | 226 |
| 22 | AChR | 71 - 80 | M | Rituximab | None | 1 | 244 | 244 |

## DISCUSSION

BCDT is one of the most impactful therapeutic advancements for the treatment of MS to date *(7)*. Nevertheless, the mechanism by which BCDTs mediate their clinical efficacy remains entirely unknown. As mentioned above, the cardinal feature of B cells, namely the production of antibodies cannot be responsible for the effect in MS. Nevertheless, it is surprising that the effect that BCDT has on disease development has not yet been revealed. Here, we sought to provide an unbiased roadmap of the systemic immune compartment in response to the elimination of B cells. Given that B cell depletion eliminates 5 to 10 % of lymphocytes from the PBMC compartment, the subtlety of the consequences for the remaining immune compartment is surprising. To better understand how BCDT alters the non-B-cell immune compartment, we designed an immunophenotyping protocol combined with a comprehensive algorithm-guided analysis.

Here, we demonstrated a selective influence of BCDTs on the CD4^+^ T cell compartment in the peripheral blood of MS patients. We observed an upregulation of surface CD27 levels along the Th cell differentiation path in response to BCDTs, that was most pronounced in memory Th cells and Tfh cells. Upon sustained signaling CD27 is cleaved into its soluble form. Therefore its upregulation in response to BCDT could be indicative of an attenuation of the CD27/CD70 signaling pathway. It is conceivable that dysregulated interactions between B and Th cells act as drivers of MS and depletion of B cells could impair these interactions. In parallel to its impairment upon BCDTs, loss of surface CD27 molecules in T cells is a molecular feature of terminal differentiation *(56, 62)*. The BCDT induced upregulation of surface CD27 may imply an impediment in the activity of Th cells, partially explaining the efficacy of the therapies.

In line with this hypothesis, increased levels of sCD27 in the CSF is a well-established biomarker of MS *(42)* and has been positively correlated to relapse rates *(63)*. Furthermore, sCD27 in the CSF of MS patients was highly predictive of active intrathecal inflammation *(43)*. Reduced sCD27 levels in the CSF has been reported in response to common therapies in MS, including BCDTs *(64–66)*. Additional support of the involvement of the CD27/CD70 pathway in MS comes from studies using preclinical models of MS. It was shown that administration of anti-CD70 monoclonal antibody significantly repressed experimental autoimmune encephalomyelitis (EAE) and Theiler’s murine encephalomyelitis virus-induced demyelinating disease (TMEV-IDD) and resulted in the reduction of clinical scores as well as inflammation and demyelination levels *(67, 68)*. Similarly, in a transgenic mouse, where the T-cell receptor recognizes the myelin oligodendrocyte glycoprotein, constitutive CD70 expression on B cells increased susceptibility of mice towards spontaneous EAE development *(69)*.

In addition to a dysregulation in the B cell/T cell interface via the CD27/CD70 axis, we observed a significant decrease in the frequency of Tfh cells in MS patients, after treatment with BCDTs. Given the central role of Tfh cells in providing costimulatory maturation and proliferation signals to B cells, their decrease in frequency after depletion of B cells highlights the importance of direct cell-cell interactions during the immunopathology of MS. Numerous studies underscore the importance of Tfh cells in MS pathophysiology *(36, 37, 70)*. CXCL13, a chemokine involved in the migration of B cells and Tfh cells *(71, 72)*, is found to be elevated in the CSF of MS patients *(73)* and predictive of a more severe disease course *(74)*. Furthermore, rituximab treatment has been shown to decrease CXCL13 as well as T cells in the CSF, in a proportional manner *(75)*. CXCL13 is prominently produced by actively demyelinating lesions but not by chronic inactive lesions *(76–78)*. Finally, increased levels of circulating Tfh cells have been reported in MS and found to be reduced in patients with complete remission *(36, 37)*.

In conclusion, our study highlights a highly selective influence of BCDTs on the Th cell compartment of MS patients. These findings suggest that BCDTs may cause a partial blockade in the full effector differentiation of Th cells due to compromised CD27/CD70 signaling. Furthermore, our analysis reports a significant reduction in the frequency of Tfh cells and emphasizes their involvement in MS as suggested by others. Our study thereby identifies a potential mechanism of action for BCDTs and reveals important insights into the pathophysiology of MS.

It was vital for this study, to include patients treated with BCDT and to obtain samples prior as well as after the treatment induction. This fact is mainly responsible for the relatively limited sample size and therefore limited statistical power. Nevertheless, our findings could be validated in patient cohorts obtained from two independent clinical centers. Current untargeted single-cell technologies (e.g. RNA-seq) are limited in their throughput and resolution, but they could have potentially revealed additional alterations driven by BCDT. As a compromise here we used CyTOF, which allowed for a combined analysis of multiple barcoded patient samples. Even though the dynamic range for detection of protein expression and reproducibility across the samples is very good, the quasi-targeted nature of CyTOF may create a bias. Finally, this study would have benefitted from access to longitudinal CSF samples, which likely are more reflective of the events that occur within inflammatory CNS lesions.

## MATERIALS AND METHODS

### Study design

The objective of this study was to investigate the immune landscape of MS patients in response to BCDTs. For this purpose longitudinal samples, before and after therapy induction, were analyzed via CyTOF. Two overlapping mass cytometry panels were applied interrogating various co-stimulatory and trafficking molecules, cytokines, and lineage markers. Data was acquired in two independent runs and normalized through batch correction. A computational pipeline comprising clustering, dimensionality reduction and statistical analyses was employed in order to retrieve phenotypic immune alterations in response to BCDT. Our findings were validated in an independent cohort from a distinct clinical center.

### Longitudinal PBMC samples of MS patients during BCDT in the discovery cohort

For the initial discovery cohort longitudinal PBMC samples of MS patients before and after BCDT were collected at the MS center of the University Hospital Basel (Table 1). MS diagnosis was based on the revised McDonald criteria*(79)*. All patients gave written informed consent, with ethical oversight by the Ethikkommission beider Basel (Ref.Nr. EK: 49/06).

In the discovery cohort samples from multiple timepoints were available. Inclusion criteria for the discovery cohort included longitudinal patient samples, appropriate B cell depletion after BCDTs as well as no previous disruptive treatments. When performing longitudinal statistical analyses, samples from timepoints closest to initial treatment were chosen. Samples containing the highest B cell frequency after treatment were chosen for the analysis of repopulating B cells and a 100-cell cut-off was applied. As such 8 patients out of 12 were qualified.

### Longitudinal PBMC samples of MS and MG patients during BCDT in the validation cohort

Cryopreserved longitudinal PBMC samples of MS and MG patients before and after BCDT in the validation cohort were collected at the Department of Neurology at Karolinska University Hospital (Table 2, Table 3). MS patients were diagnosed according to the revised McDonald criteria*(79)*. The MG diagnosis was confirmed by at least 2 of the following: a positive AChR antibody test result, an abnormal electrophysiological test result (repetitive nerve stimulation and/or single fiber electromyography) consistent with MG, and/or a clinically significant response to an oral or intravenous AChEI test (per treating physician’s opinion). All patients had given written informed consent and the study has been approved by the regional ethical review board of Stockholm (MS dnr. 2009/2107-21/2, last amendment dnr. 2022-03650-02; MG 2016-827-31).

In the validation cohort each patient had one timepoint before treatment and one timepoint after treatment. Inclusion criteria for the validation cohort included appropriate B cell depletion after BCDTs.

### Statistical analysis

All statistical analyses were done in R. Wilcoxon effect size calculations were done with the R package “rstatix” and were used in the analysis of marker changes in the replenished B cell compartment. Non-parametric paired Wilcoxon rank-sum test from the R package “stats” was used to compare two groups unless stated otherwise. Controlling for multiple comparisons was accomplished with the Benjamini-Hochberg approach also from the “stats” package.

### Study approval

All donors had given written informed consent and the study was approved by the regional ethical review board of Stockholm (MS dnr. 2009/2107-21/2, last amendment dnr. 2022-03650-02; MG 2016-827-31) and Basel (Ref.Nr. EK: 49/06), respectively.

## Data Availability

Mass cytometry data of the discovery and validation cohort will be made publicly available upon publication. Custom analysis code to reproduce the analysis will be uploaded to a public repository.

## SUPPLEMENTARY MATERIALS

Materials and Methods

Figs. S1 and S2

Tables S1 to S3

References (80–86)

## Acknowledgments

We thank all the patients that participated in the study. We thank Catarina Raposo for the helpful review of the manuscript.

## Author contributions

E.G. and F.I. generated the mass cytometry data. C.U. analyzed all the data. I.C., M.K., F.P., N.S., R.F., T.O. and T.D. performed clinical characterization of the cohorts and provided patient samples and clinical input. B.B. and F.I. jointly supervised the study. B.B. funded the study. C.U., F.I. and B.B. wrote the manuscript.

