## Supplement for "B cell depletion attenuates CD27 signaling of T helper cells in multiple sclerosis"

Ulutekin *et al.*

**This Word document includes:**

Materials and Methods

Figs. S1 and S2

Tables S1 – S3

References (80–86)

### Supplementary Materials

#### Materials and Methods

##### Ex vivo activation of PBMCs from MS patients

For intracellular cytokine detection, PBMCs were activated as described previously *(80)*. In brief, PBMCs were stimulated with 50 ng/ml phorbol 12-myristate 13-acetate (Sigma-Aldrich), 500 ng/ml ionomycin (Sigma-Aldrich), 1× brefeldin A (BD Biosciences) and 1× monensin (BD Biosciences) in cell culture medium (RPMI-1640, 10% FCS (Biochrom), 1× L-glutamine (Life Technologies) and 1× penicillin–streptomycin (Life Technologies)) at 37°C for 4h. PBMCs were washed once in cell culture medium and barcoded subsequently.

##### Live cell barcoding for mass cytometry

PBMCs of both, the discovery, and the validation cohort, were processed and acquired in two independent batches each. Each batch contained PBMCs of a healthy donor that was used as normalization control to quantify batch effects between individual experiments. Longitudinal samples of the same patient were acquired in the same batch. Two panels were used to acquire the samples from the discovery cohort, one panel directed against surface epitopes of unmanipulated PBMCs and one panel to detect intracellular cytokines after *ex vivo* activation. The intracellular cytokine panel after *ex vivo* activation was used to acquire the samples from the validation cohort. The full list of antigens per panel, clones of antibodies utilized, and heavy metal tags are listed in table S1 to S3.

PBMCs of each individual batch were barcoded using a restricted live cell barcoding strategy as described before *(81)*. In short, anti-CD45 monoclonal antibodies were tagged with $^{89}$Y, $^{104}$Pd, $^{105}$Pd, $^{106}$Pd, $^{108}$Pd, $^{110}$Pd, $^{115}$In and $^{181}$Ta isotopes and a restricted 8-choose-3 barcoding scheme was applied (table S1 to S3). PBMCs of each sample were labelled with a unique barcode consisting of three different heavy metal-tagged anti-CD45 antibodies in RPMI-1640 and 4% FCS for 25 min. Barcoded cells were washed twice with RPMI-1640 and 4% FCS and pooled into one single reaction vessel.

##### Surface and intracellular cytokine staining for mass cytometry

The barcoded sample convolute was labelled with a mix of antibodies directed against surface epitopes in RPMI-1640 and 4% FCS for 40 min at 37°C. Live/dead discrimination was achieved by incubating the sample convolute in 2.5 µM cisplatin (Standard BioTools Inc.) in PBS for two minutes on ice followed by the addition of 2% FCS in PBS for two more minutes.

For the antibody panel directed against surface epitopes of unmanipulated PBMCs, cells were fixed in 1X FOXP3 fixation/permeabilization buffer (BioLegend) for 40 min at 4°C to facilitate transcription factor labelling. Cells were washed twice in with permeabilization buffer (0.5% saponin (Sigma-Aldrich), 2% BSA (Sigma-Aldrich), 0.01% sodium azide (Sigma-Aldrich) in PBS) and labelled with antibody mix in permeabilization buffer for 1 h at 4°C.

For the antibody panel directed against intracellular cytokines, cells were fixed in 1.6% PFA (Electron Microscopy Sciences) for 1h at 4°C washed with permeabilization buffer. Cells were incubated with the antibody mix directed against cytokines in permeabilization buffer for 1 h at 4°C.

For both panels, PBMCs were washed in PBS and incubated in 1X iridium intercalator solution (Standard BioTools Inc.) at 4°C overnight. Cells were washed twice with PBS and twice with MaxPar water (Standard BioTools Inc.).

#### Mass cytometry data acquisition and preprocessing

The barcoded sample convolute for each panel and cohort was acquired in two independent batches. Each batch was acquired on a CyTOF 2.1 (Standard BioTools Inc.) and instrument performance and tuning was checked on a daily basis. Data were normalized using five-element beads (Standard BioTools Inc.) *(82)*. Live singlets were identified by manual gating based on the parameters event length, center, width, DNA content ($^{191/193}Ir$) and live/dead staining ($^{195}Pt$) in FlowJo (BD Biosciences). Living cells in the sample convolute were debarcoded by applying manual Boolean gates in FlowJo (BD Biosciences). Only cells bearing exactly three distinct metal barcodes were extracted to exclude doublets and prevent misidentification of barcodes. Data of debarcoded samples were transformed in R, using a hyperbolic arcsin function with variable cofactors (5-300). The data was normalized between 0 and 1 using the 99.95$^{th}$ percentile.

#### Batch Normalization

A batch effect was observed in the data obtained using the cytokine panel across two independent acquisitions. Longitudinal samples of patients were present within the same batch, as such, this batch effect was most impactful during clustering, generating batch specific clusters. To minimize the effect, batch normalization was done using the R package "CytoNorm" *(83)*. The training and application of the algorithm was done just after the transformation of the data. Healthy donor samples present in both batches were used to train the algorithm, and no marker was excluded from normalization. The normalization was quality tested using biaxial plots sequentially checking for all clusters, to ensure there was no splitting or merging of valid populations.

#### High-Dimensional Analysis

The data was initially clustered using the R package "FlowSOM" *(84)* and generated 100 clusters based on the expression of all available markers except for cytokines. The "ConsensusClusterPlus" *(85)* package was used subsequently to sequentially merge the cluster until 30 clusters were achieved. To identify biologically meaningful cell types, the final clusters were merged manually and annotated based on marker expression characteristics. The R package "umap" *(86)* was used to generate the UMAP coordinates. The R packages "ComplexHeatmap" and "ggplot2" were used for the visualizations of the data in various plots. The R packages "flowCore", "dplyr", "tidyr" and "tibble" were used for loading and reformatting the data.

#### Supplementary Figures

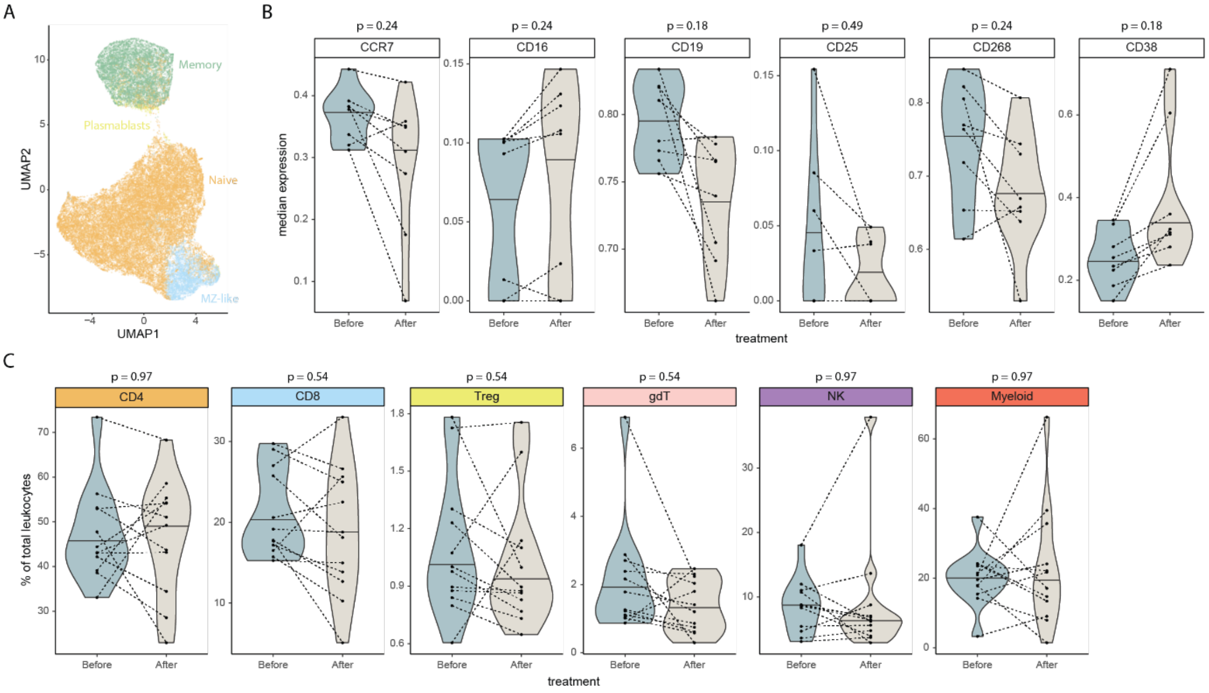


**Fig. S1. B cell reconstitution and main cell frequency alterations.** (**A**) UMAP of 50,000 cells randomly sampled from the data. Color code indicated the B cell sub-clusters generated through clustering and manual annotation. (**B**) Violin plots showing changes in select marker expression naïve B cells in BCDT treated MS. P values correspond to the paired Wilcoxon rank-sum test applying a Benjamini-Hochberg correction. (**C**) Violin plots showing changes in the frequency across main cell clusters in BCDT treated MS. Paired Wilcoxon rank-sum test and Benjamini-Hochberg correction was applied.

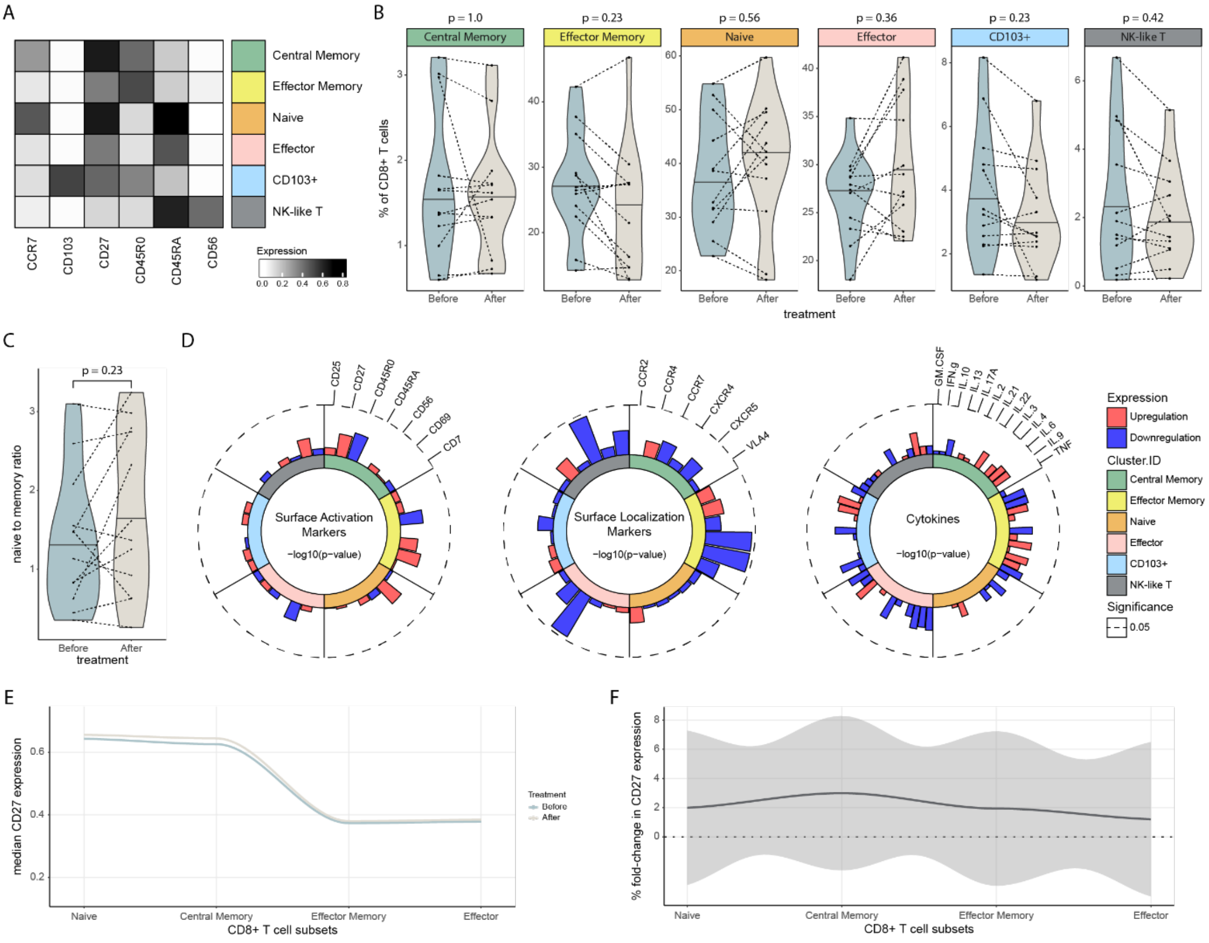

**Fig. S2. BCDTs does not affect the peripheral CD8$^{+}$ T cell landscape of MS patients.** (**A**) Heatmap showing median expression of lineage markers across identified CD8$^{+}$ T cell subsets. (**B**) Violin plots showing changes in the frequency across CD8+ T cell subsets in BCDT treated MS. Paired Wilcoxon rank-sum test and Benjamini-Hochberg correction was applied. (**C**) Violin plots showing changes in the naive to memory ratio within the CD8$^{+}$ T cell compartment. Paired Wilcoxon rank-sum test was applied. (**D**) Radar plots showing changes in the marker expression of CD8$^{+}$ T cell subsets in BCDT treated MS. Values correspond to -log10(p-value) and were adjusted with the Benjamini-Hochberg correction. Inner circle color annotations denote each respective subset. Bar color denotes the upregulation or downregulation of the respective marker. Marker order is consistent across subsets. (**E**) Median CD27 expression along CD8$^{+}$ T cell subsets of the MS patients. Points are individual sample values. Color code denotes patient values before or after treatment. (**F**) Percent fold-change in CD27 expression along CD8$^{+}$ T cell subsets of the MS patients, depicted in percent changes. Shaded area represents the 95 % confidence interval.

#### Supplementary Tables

**Table S1:** Discovery cohort, cytokine panel.

| Isotype | Metal | Antigen | Clone | Supplier | Category | Batch |
|---|---|---|---|---|---|---|
| 141 | Pr | VLA4 | 9F10 | Fluidigm | surface | 1 + 2 |
| 142 | Nd | CD19 | HIB19 | Fluidigm | surface | 1 + 2 |
| 143 | Nd | CCR2 | K036C2 | Biolegend | surface | 1 + 2 |
| 144 | Nd | IL.4 | MP4-25D2 | Biolegend | ICS | 1 + 2 |
| 145 | Nd | CD4 | RPA-T4 | Fluidigm | surface | 1 + 2 |
| 146 | Nd | CD8 | RPA-T8 | Fluidigm | surface | 1 + 2 |
| 147 | Sm | IL.2 | MQ1-17H12 | Biolegend | ICS | 1 + 2 |
| 148 | Nd | IL.17A | BL168 | Fluidigm | ICS | 1 + 2 |
| 149 | Sm | IL.3 | BVD8-3G11 | Biolegend | ICS | 1 + 2 |
| 150 | Nd | IL.22 | 22URTI | Fluidigm | ICS | 1 + 2 |
| 151 | Eu | CD103 | Ber-ACT8 | Fluidigm | surface | 1 + 2 |
| 152 | Sm | TCRgd | 11F2 | Fluidigm | surface | 1 + 2 |
| 153 | Eu | CD25 | M-A251 | Biolegend | surface | 1 + 2 |
| 154 | Sm | IL.6 | MQ2-13A5 | Biolegend | ICS | 1 + 2 |
| 155 | Gd | IL.9 | MH9A4 | Biolegend | ICS | 1 + 2 |
| 156 | Gd | IL.13 | JES10-5A2 | Biolegend | ICS | 1 + 2 |
| 158 | Gd | CCR4 | 205410 | Fluidigm | surface | 1 + 2 |
| 159 | Tb | GM.CSF | BVD2-21C11 | Fluidigm | ICS | 1 + 2 |
| 160 | Gd | CD69 | FN50 | Biolegend | surface | 1 + 2 |
| 161 | Dy | CD20 | 2H7 | Biolegend | surface | 1 + 2 |
| 162 | Dy | CD27 | O323 | Biolegend | surface | 1 + 2 |
| 163 | Dy | CD7 | 6B7 | Biolegend | surface | 1 + 2 |
| 164 | Dy | CD45R0 | UCHL1 | Fluidigm | surface | 1 + 2 |
| 165 | Ho | IFN.g | B27 | Fluidigm | ICS | 1 + 2 |
| 166 | Er | IL.10 | JES3-9D7 | Fluidigm | ICS | 1 + 2 |
| 167 | Er | CCR7 | G043H7 | Fluidigm | surface | 1 + 2 |
| 168 | Er | TNF | MAb11 | Biolegend | ICS | 1 + 2 |
| 169 | Tm | CD45RA | HI100 | Fluidigm | surface | 1 + 2 |
| 170 | Er | CD3 | UCHT-1 | Fluidigm | surface | 1 + 2 |
| 171 | Yb | CXCR5 | 51505 | Fluidigm | surface | 1 + 2 |
| 172 | Yb | IL.21 | 3A3-N2 | Fluidigm | ICS | 1 + 2 |
| 173 | Yb | CXCR4 | 12G5 | Fluidigm | surface | 1 + 2 |
| 174 | Yb | HLADR | L243 | Fluidigm | surface | 1 + 2 |
| 175 | Lu | CD14 | M5E2 | Fluidigm | surface | 1 + 2 |
| 176 | Yb | CD56 | NCAM16.2 | Biolegend | surface | 1 + 2 |
| 209 | Bi | CD16 | 3G8 | Fluidigm | surface | 1 + 2 |

**Table S2:** Validation cohort, cytokine panel.

| Isotype | Metal | Antigen | Clone | Supplier | Category | Batch |
|---|---|---|---|---|---|---|
| 141 | Pr | VLA4 | 9F10 | Fluidigm | surface | 1 + 2 |
| 142 | Nd | CD19 | HIB19 | Fluidigm | surface | 1 + 2 |
| 143 | Nd | CCR2 | K036C2 | Biolegend | surface | 1 + 2 |
| 144 | Nd | IL.4 | MP4-25D2 | Biolegend | ICS | 1 + 2 |
| 145 | Nd | CD4 | RPA-T4 | Fluidigm | surface | 1 + 2 |
| 146 | Nd | CD8 | RPA-T8 | Fluidigm | surface | 1 + 2 |
| 147 | Sm | IL.2 | MQ1-17H12 | Biolegend | ICS | 1 + 2 |
| 148 | Nd | IL.17A | BL168 | Fluidigm | ICS | 1 + 2 |
| 149 | Sm | IL.3 | BVD8-3G11 | Biolegend | ICS | 1 + 2 |
| 150 | Nd | IL.22 | 22URTI | Fluidigm | ICS | 1 + 2 |
| 151 | Eu | CD103 | Ber-ACT8 | Fluidigm | surface | 1 + 2 |
| 152 | Sm | TCRgd | 11F2 | Fluidigm | surface | 1 + 2 |
| 153 | Eu | CD25 | M-A251 | Biolegend | surface | 1 + 2 |
| 154 | Sm | IL.6 | MQ2-13A5 | Biolegend | ICS | 1 |
| 155 | Gd | IL.9 | MH9A4 | Biolegend | ICS | 1 + 2 |
| 156 | Gd | IL.13 | JES10-5A2 | Biolegend | ICS | 1 + 2 |
| 158 | Gd | CCR4 | 205410 | Fluidigm | surface | 1 + 2 |
| 159 | Tb | GM.CSF | BVD2-21C11 | Fluidigm | ICS | 1 + 2 |
| 160 | Gd | CD69 | FN50 | Biolegend | surface | 1 + 2 |
| 161 | Dy | CD20 | 2H7 | Biolegend | surface | 1 + 2 |
| 162 | Dy | CD27 | O323 | Biolegend | surface | 1 + 2 |
| 163 | Dy | CD7 | 6B7 | Biolegend | surface | 1 + 2 |
| 164 | Dy | CD45R0 | UCHL1 | Fluidigm | surface | 1 + 2 |
| 165 | Ho | IFN.g | B27 | Fluidigm | ICS | 1 + 2 |
| 166 | Er | IL.10 | JES3-9D7 | Fluidigm | ICS | 1 + 2 |
| 167 | Er | CCR7 | G043H7 | Fluidigm | surface | 1 + 2 |
| 168 | Er | TNF | MAb11 | Biolegend | ICS | 1 + 2 |
| 169 | Tm | CD45RA | HI100 | Fluidigm | surface | 1 + 2 |
| 170 | Er | CD3 | UCHT-1 | Fluidigm | surface | 1 + 2 |
| 171 | Yb | CXCR5 | 51505 | Fluidigm | surface | 1 + 2 |
| 172 | Yb | IL.21 | 3A3-N2 | Fluidigm | ICS | 1 + 2 |
| 173 | Yb | CXCR4 | 12G5 | Fluidigm | surface | 1 + 2 |
| 174 | Yb | HLADR | L243 | Fluidigm | surface | 1 |
| 175 | Lu | CD14 | M5E2 | Fluidigm | surface | 1 + 2 |
| 176 | Yb | CD56 | NCAM16.2 | Biolegend | surface | 1 + 2 |
| 209 | Bi | CD16 | 3G8 | Fluidigm | surface | 1 + 2 |
| 174 | Yb | CD28 | CD28.2 | Biolegend | surface | 2 |

**Table S3:** Discovery cohort, surface panel.

| Isotype | Metal | Antigen | Clone | Supplier | Category | Batch |
|---|---|---|---|---|---|---|
| 141 | Pr | CCR6 | G034E3 | Fluidigm | surface | 1 + 2 |
| 142 | Nd | CD19 | HIB19 | Fluidigm | surface | 1 + 2 |
| 143 | Nd | CCR2 | K036C2 | Biolegend | surface | 1 + 2 |
| 144 | Nd | CCR5 | J418F1 | Biolegend | surface | 1 + 2 |
| 145 | Nd | CD4 | RPA-T4 | Fluidigm | surface | 1 + 2 |
| 146 | Nd | CD8 | RPA-T8 | Fluidigm | surface | 1 + 2 |
| 147 | Sm | CD11c | BU15 | Biolegend | surface | 1 + 2 |
| 148 | Nd | CD16 | 3G8 | Fluidigm | surface | 1 + 2 |
| 149 | Sm | CD25 | 2A3 | Biolegend | surface | 1 + 2 |
| 150 | Nd | CD27 | LG.3A10 | Fluidigm | surface | 1 + 2 |
| 151 | Eu | CD38 | HIT2 | Biolegend | surface | 1 + 2 |
| 152 | Sm | TCRgd | 11F2 | Fluidigm | surface | 1 + 2 |
| 153 | Eu | CD45RA | HI100 | Fluidigm | surface | 1 + 2 |
| 154 | Sm | CD3 | UCHT1 | Fluidigm | surface | 1 + 2 |
| 155 | Gd | CD268 | 11C1 | Fluidigm | surface | 1 + 2 |
| 156 | Gd | CXCR3 | G025H7 | Fluidigm | surface | 1 + 2 |
| 158 | Gd | CCR4 | 205410 | Fluidigm | surface | 1 + 2 |
| 159 | Tb | CD116 | 4H1 | Biolegend | surface | 1 + 2 |
| 160 | Gd | CD69 | FN50 | Biolegend | surface | 1 + 2 |
| 161 | Dy | CD20 | 2H7 | Biolegend | surface | 1 + 2 |
| 162 | Dy | FOXP3 | PCH101 | Fluidigm | transcription factor | 1 + 2 |
| 163 | Dy | CD7 | 6B7 | Biolegend | surface | 1 + 2 |
| 164 | Dy | CD45R0 | UCHL1 | Fluidigm | surface | 1 + 2 |
| 165 | Ho | CD127 | A019D5 | Fluidigm | surface | 1 + 2 |
| 166 | Er | CD86 | IT.2 | Biolegend | surface | 1 + 2 |
| 167 | Er | CCR7 | G043H7 | Fluidigm | surface | 1 + 2 |
| 168 | Er | Ki67 | B56 | Fluidigm | transcription factor | 1 + 2 |
| 169 | Tm | CD33 | WM53 | Fluidigm | surface | 1 + 2 |
| 170 | Er | IgD | IA6-2 | Biolegend | surface | 1 + 2 |
| 171 | Yb | CD22 | HIB22 | Biolegend | surface | 1 + 2 |
| 172 | Yb | IgM | MHM-88 | Fluidigm | surface | 1 + 2 |
| 173 | Yb | CD56 | NCAM16.2 | Biolegend | surface | 1 + 2 |
| 174 | Yb | HLADR | L243 | Fluidigm | surface | 1 + 2 |
| 175 | Lu | CD14 | M5E2 | Fluidigm | surface | 1 + 2 |
| 176 | Yb | CD5 | UCHT2 | Biolegend | surface | 1 + 2 |
| 209 | Bi | CD11b | ICRF44 | Fluidigm | surface | 1 + 2 |
